# What most influences severity and death of COVID-19 patients in Brazil? Is it clinical, social, or demographic factors? An observational study

**DOI:** 10.1101/2021.06.03.21258128

**Authors:** Ana Carolina C. N. Mafra, Régis Rodrigues Vieira, Camila N. Monteiro, Denise F. B. Cavalcante, João L. Miraglia, Daiana Bonfim, Danielle C. Palacio, Alessandra C. F. Martins, Letícia Yamawaka de Almeida, João Peres Neto

## Abstract

**Objective:** This study aimed to assess the space distribution and factors associated with the risk of severe acute respiratory syndrome (SARS) and death in COVID-19 patients, based on routine register data; and to develop and validate a predictive model of the risk of death from COVID-19.

**Methods:** A cross-sectional, epidemiological study of positive SARS-CoV-2 cases, reported in the south region of the city of São Paulo, SP, Brazil, from March 2020 to February 2021. Data were obtained from the official reporting databases of the Brazilian Ministry of Health for influenza-like illness (ILI) (esus-VE, in Portuguese) and for patients hospitalized for SARS (SIVEP-Gripe). The space distribution of cases is described by 2D kernel density. To assess potential factors associated with the outcomes of interest, generalized linear and additive logistic models were adjusted. To evaluate the discriminatory power of each variable studied as well as the final model, C-statistic was used (area under the receiver operating characteristics curve). Moreover, a predictive model for risk of death was developed and validated with accuracy measurements in the development, internal and temporal (March and April 2021) validation samples.

**Results:** A total of 16,061 patients with confirmed COVID-19 were enrolled. Morbidities associated with a higher risk of SARS were obesity (OR=25.32) and immunodepression (OR=12.15). Morbidities associated with a higher risk of death were renal disease (OR=11.8) and obesity (OR=8.49), and clinical and demographic data were more important than the territory *per se*. Based on the data, a calculator was developed to predict the risk of death from COVID-19, with 92.2% accuracy in the development sample, 92.3% in the internal validation sample, and 80.0% in the temporal validation sample.

**Conclusions:** The risk factors for SARS and death in COVID-19 patients seeking health care, in order of relevance, were age, comorbidities, and socioeconomic factors, considering each discriminatory power.

## Introduction

In December 2019, the first cases of SARS-CoV-2 infection were described in Wuhan, China, and 16 months later COVID-19 has affected approximately 140 million people and resulted in more than 3 million deaths worldwide [1].

The American continent is currently the region most affected by the COVID-19 pandemic, and Latin America has one of the highest COVID-19 mortality rates in the world, driven by the inequality within its countries. Brazil has more than 200 million inhabitants and is a country marked by social inequality. Up to April 2021, SARS-CoV-2 has infected more than 14 million people and caused over 382 thousand deaths. São Paulo is the largest city in the country, with 12 million inhabitants, and more than 980 thousand have been infected and 26 thousand have died [1,2].

The country has a mixed public and private health system, with one of the largest public health care programs in the world; in 2020, Brazil’s Unified Health Care System (SUS) completed 30 years of existence. Approximately 75% of the Brazilian population depends exclusively on the public health care system. Despite the advances following expansion of health services, particularly in Primary Health Care (PHC), SUS has suffered with chronic underfunding, resource scarcity and deficient delivery of services [3].

In the last five years, Brazil has undergone structural changes leading to more severe inequalities, which have strongly impacted the response to the pandemic. The following changes stand out: the spending ceiling that has frozen health care financing starting in 2016 through the next 20 years; changes in the National Primary Care Policy, in 2017, leading to uncertainty with flexibilization of the need for community health workers, and weakening of the Family Health Strategy, one of the pillars of PHC in the country. Additionally, in 2019 the program “Previne Brasil” [Prevent Brazil] was launched and changed the PHC funding scenario, increasing doubts about the ability of SUS to keep its universal nature, at the risk of reducing PHC to a selective model, based on focal targets [4].

The risk factors strongly associated with the severity of COVID-19 are known, particularly advanced age, cancer, chronic renal disease, chronic obstructive pulmonary disease, cardiovascular disease, prior transplantation, obesity, pregnancy, sickle cell anemia, smoking and diabetes mellitus [5–8]. However, there is no specific data on risk factors associated with high mortality rates in territories of low-income and household overcrowding, such as Brazilian slums [9].

The combination of the SARS-CoV-2 epidemic with chronic non-communicable diseases (NCD) in a social context of poverty and inequality has disproportionally affected the so-called slums, i.e. densely populated and poverty-stricken areas [10].

The territorial distribution of COVID-19 in Brazil started in large, more populous urban centers, where trade relations are more intense, and flowed along the richest transport routes in the country, shaping virus spread, and ultimately reaching poorer regions [11]. São Paulo is one of the cities with the highest incidence of cases, mostly driven by its high population density. However, there is no specific data on how virus transmission occurred within the most vulnerable areas of the city [12], such as the districts of Vila Andrade and Campo Limpo, regions stricken by great social inequality and extreme vulnerability.

Despite the known social inequalities among countries and even regions within the same country, and the need for differentiated responses in highly vulnerable and resource-scarce settings, little is known about the specific realities of virus spread and the risk factors present in slums. The data extrapolated is from regions with vastly different conditions, which may not correspond to the real world [11]. Hence, this study aimed to assess the space distribution and factors associated with the risk of SARS and death from COVID-19, based on routinely collected register data, as well as to develop and validate a predictive model of risk of death from COVID-19.

## Methods

This was a cross-sectional, epidemiological study that included positive COVID-19 cases reported in the south region of the city of São Paulo, SP, Brazil, between March 2020 and February 2021. We conduct the work following the STROBE checklist (S2 Text).

The study territory includes two administrative districts, Campo Limpo and Vila Andrade, which together have an estimated population of approximately 400 thousand people (Fig 1A) [13]. In this region, social vulnerability can be measured by the São Paulo Social Vulnerability Index (IPVS, in Portuguese) [14], which ranges from 1 for extremely low vulnerability (navy areas) to 6 for very high vulnerability (purple areas) (Fig 1B). The GeoSES is another socioeconomic indicator [15] that can express the region’s vulnerability. It is a composite index that summarizes the Brazilian socioeconomic context, and is composed by seven dimensions: education, mobility, poverty, wealth, income, segregation and deprivation of resources and services. The index can range from −1 (worst socioeconomic context) to 1 (best socioeconomic context). The districts of Vila Andrade and Campo Limpo have average values of −0.03 and −0.36 respectively (Fig 1C).

**Fig 1.**
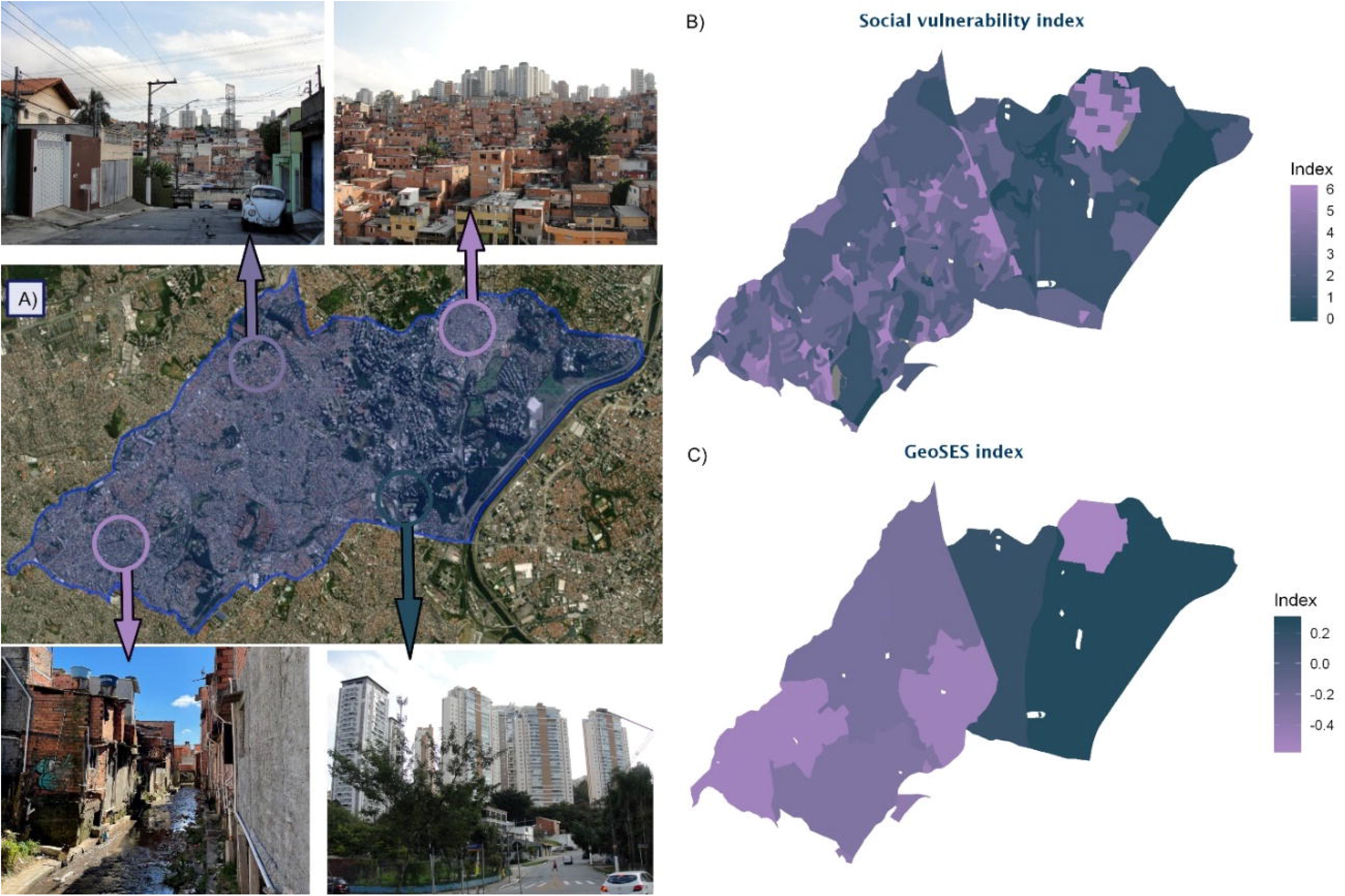
Study territory. A: Satellite image and photographs of the region. B: Distribution of the São Paulo Social Vulnerability Index. C: Distribution of the GeoSES Socioeconomic Index.

The distribution of the health care facilities, privet and public, accessed by the study population can be visualized in a map available in S1 Fig. Health care facilities were categorized as privet or public according to the National Registry of Health Care Facilities (CNES, in Portuguese), and was considered in the analyses as an indicator of health care access, since public health services are universally accessible, while only those with health insurance or high income have access to private services.

The data reviewed come from official reporting databases of the Brazilian Ministry of Health, including two systems: one for influenza-like illness (esus-VE), with milder cases of the disease reported by any health care facility, and another (SIVEP-Gripe) for patients hospitalized with severe acute respiratory syndrome (SARS). The residence location of individuals included in the study was geocoded by GISA/CEINFO - SMS, and made available by the Health Surveillance Unit of Campo Limpo. The study database was anonymized, with all identifiable patient information removed. This study was approved by the Institutional Review Boards of Hospital Israelita Albert Einstein and the São Paulo Health Department, protocol numbers 4.462.994 and 4.648.956, respectively.

The study population was defined as all COVID-19 cases confirmed by PCR. The outcome variables were progression to SARS and death, categorized as: influenza-like illness (ILI)/severe acute respiratory syndrome (SARS) and discharge/death, respectively.

The independent variables were sociodemographic characteristics, comorbidities, and symptoms. The following inclusion criteria were used: to be a resident of the study region, aged 18 years and older, with laboratory confirmation of COVID-19 by PCR. When the same patient had multiple records, and ILI had been reported previously to SARS, only the SARS record was considered (Fig 2).

**Fig 2.**
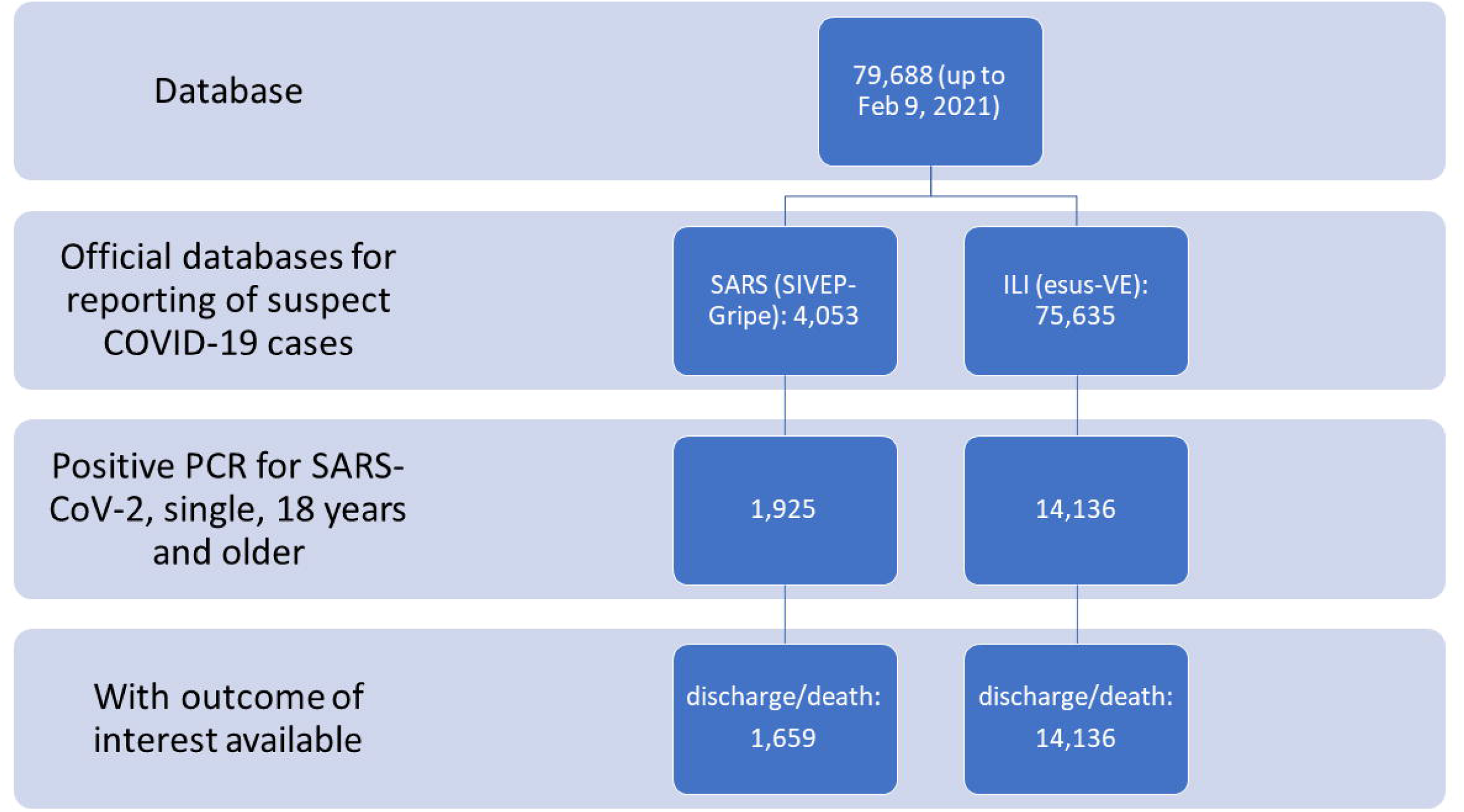
Study data inclusion and exclusion flow.

Results were presented as absolute and relative frequencies, overall and by outcome (death or SARS). For simple comparison between outcome groups, the chi-square test was used for categorical variables and Student’s t-test was used for numerical variables.

The spatial distribution of cases was described by a 2D kernel density.

To assess potential factors associated with the outcomes of interest, generalized linear and additive logistic models were adjusted, and the latter were used for inclusion of the spatial location of subjects’ place of residence (latitude and longitude). First, simple models were adjusted, and variables with a p value ≤0.2, and with less than 10% of missing values were considered for the multiple models. No data imputation was used, so only complete observations were used in the analyses. Symptoms were not included in the model, so it would only evaluate conditions that existed prior to COVID-19. The selection of variables for the final model was performed by backwards stepwise elimination, with a p value <0.05 as the criteria to inclusion in the model. In the final model, the quality of adjustment was verified by the magnitude of standard errors, the influence of inclusion or exclusion of variables in the estimated odds ratio, and the variance inflation factor, which was not greater than 1.2.

To assess the discriminatory power of each variable evaluated as well as the final model, the C-statistic was used (area under the receiver operating characteristics curve).

Since the multiple model presented a good discriminatory power, a predictive model for death was developed, with the main clinical and demographic characteristics of patients arriving at health care facilities for COVID-19 evaluation. In this analysis, the symptoms were also considered, and we followed the same method of variable selection presented above. The data was randomly split in two sets with a 2:1 ratio for training and testing, respectively. The predictive performance of the model was assessed by accuracy measurements, discriminatory power and the Hosmer-Lemeshow test [16]. The temporal validation used data from patients over 18 years of age, with a positive COVID-19 test and who had symptoms onset between March and April of 2021.

All analyses were performed with the R software, version 3.6.3 [17], with the mgcv [18], ggplot2 [19] and viridis [20] packages. A 5% significance level was adopted.

## Results

The study investigated a population of 16,061 patients with confirmed COVID-19, between March 2020 and February 2021, with 1,925 (11.98%) cases of SARS and 375 (2.37%) deaths.

The age of included individuals ranged from 18 to 103 years, with a mean of 42.1 years (standard deviation 14.9) and a median of 40 years.

The baseline characteristics of study participants, stratified by outcome, and the C-statistic for each variable and outcome can be found at Table 1.

**Table 1.** **Baseline characteristics of study participants, overall and stratified by study outcome, and C-statistics for each variable and outcome. N=16**,**031 confirmed for COVID-19**.

|  | Overall | Missing<br>rate (%) | SARS (yes) | C SARS | Death (yes) | C<br>Death |
| --- | --- | --- | --- | --- | --- | --- |
| N | 16,061 |  | 1,925 |  | 375 |  |
| Profile |  |  |  |  |  |  |
| Mean age (SD) | 42.10 (14.94) | 0.0 | 56.93 (16.77)** | 0.78 | 68.08 (14.48)** | 0.90 |
| **Age Range** |  | 0.0 |  | 0.75 |  | 0.86 |
| 18 to 40 years | 7892 (49.1) |  | 337 (17.5)** |  | 18 (4.8)** |  |
| 40 to 60 years | 6022 (37.5) |  | 736 (38.2) |  | 82 (21.9) |  |
| 60 to 80 years | 1830 (11.4) |  | 653 (33.9) |  | 184 (49.1) |  |
| 80 years and older | 317 (2.0) |  | 199 (10.3) |  | 91 (24.3) |  |
| **Sex** |  | 0.0 |  | 0.56 |  | 0.57 |
| Female | 8804 (54.8) |  | 852 (44.3)** |  | 153 (40.8)** |  |
| Male | 7230 (45.0) |  | 1073 (55.7) |  | 222 (59.2) |  |
| Undefined | 27 (0.2) |  | 0 (0.0) |  | 0 (0.0) |  |
| **Race/skin color** |  | 24.9 |  | 0.54 |  | 0.53 |
| White | 5925 (49.1) |  | 675 (51.4)** |  | 144 (49.3) |  |
| Black | 663 (5.5) |  | 63 (4.8) |  | 21 (7.2) |  |
| Yellow | 901 (7.5) |  | 35 (2.7) |  | 10 (3.4) |  |
| Brown | 4563 (37.8) |  | 539 (41.1) |  | 117 (40.1) |  |
| Indigenous | 15 (0.1) |  | 1 (0.1) |  | 0 (0.0) |  |
| **Caucasian** |  | 24.9 |  | 0.51 |  | 0.50 |
| No | 6142 (50.9) |  | 638 (48.6) |  | 148 (50.7) |  |
| Yes | 5925 (49.1) |  | 675 (51.4) |  | 144 (49.3) |  |
| **Conditions** |  |  |  |  |  |  |
| Diabetes (y) | 997 (6.5) | 5.0 | 506 (44.9)** | 0.71 | 154 (49.4)** | 0.72 |
| Obesity (y) | 187 (1.2) | 5.0 | 118 (10.5)** | 0.55 | 44 (14.1)** | 0.56 |
| Immunodepression (y) | 145 (0.9) | 5.0 | 62 (5.5)** | 0.52 | 26 (8.3)** | 0.53 |
| Heart disease (y) | 1494 (9.8) | 5.0 | 720 (63.8)** | 0.79 | 215 (68.9)** | 0.80 |
| Renal disease (y) | 122 (0.8) | 5.0 | 85 (7.5)** | 0.54 | 53 (17.0)** | 0.58 |
| Respiratory Disease (y) | 418 (2.7) | 5.0 | 125 (11.1)** | 0.55 | 46 (14.7)** | 0.56 |
| Pregnancy (y) | 78 (0.5) | 7.9 | 11 (1.7)** | 0.51 | 0 (0.0) | 0.50 |
| **Symptoms** |  |  |  |  |  |  |
| Fever | 6868 (43.8) | 2.3 | 1211 (78.0)** | 0.69 | 235 (73.2)** | 0.65 |
| Cough | 8354 (53.0) | 1.9 | 1352 (83.3)** | 0.67 | 271 (81.9)** | 0.64 |
| Sore throat | 3755 (24.6) | 5.1 | 246 (22.3) | 0.51 | 55 (20.3) | 0.52 |
| Dyspnea | 3567 (22.7) | 2.0 | 1332 (82.7)** | 0.83 | 293 (85.4)** | 0.82 |
| Diarrhea | 231 (1.5) | 5.2 | 231 (21.1)** | 0.61 | 65 (23.6)** | 0.61 |
| Vomiting | 125 (0.8) | 5.5 | 125 (12.0)** | 0.56 | 32 (12.5)** | 0.55 |
| Abdominal pain | 43 (0.3) | 9.5 | 43 (10.8)** | 0.55 | 7 (7.7)** | 0.53 |
| Fatigue | 157 (1.1) | 9.2 | 157 (35.4)** | 0.68 | 18 (18.8)** | 0.58 |
| Loss of smell | 1487 (10.2) | 9.1 | 125 (27.3)** | 0.59 | 12 (12.5) | 0.51 |
| Loss of taste | 1386 (9.5) | 9.4 | 96 (22.8)** | 0.57 | 11 (12.1) | 0.51 |
| **Socioeconomic** |  |  |  |  |  |  |
| **Type of healthcare facility** |  |  |  |  |  |  |
| Private | 6999 (51.6) | 15.5 | 1079 (58.3)** | 0.54 | 161 (44.7)* | 0.53 |
| Public | 6570 (48.4) |  | 772 (41.7) |  | 199 (55.3) |  |
| **Mean IPVS (SD)** | 2.68 (1.60) | 0.9 | 2.57 (1.54)* | 0.52 | 2.78 (1.52) | 0.53 |
| 0-3 | 12619 (79.3) | 0.9 | 1555 (81.2)* | 0.51 | 287 (76.7) | 0.51 |
| 4-6 | 3302 (20.7) |  | 360 (18.8) |  | 87 (23.3) |  |
| **Mean GeoSES (SD)** | -0.20 (0.30) | 0.0 | -0.19 (0.30)* | 0.52 | -0.25 (0.28)* | 0.54 |
| <0 | 11438 (71.2) | 0.0 | 1336 (69.4) | 0.51 | 289 (77.1)* | 0.52 |
| >=0 | 4623 (28.8) |  | 589 (30.6) |  | 86 (22.9) |  |

C: C-statistic, representing the area under the ROC curve. Measures reported as absolute frequency
(%), when mean not indicated (standard deviation). \* p value for hypothesis testing <0.05. \*\* p
value for hypothesis testing <0.001. Tests performed: chi-square and Student's *t*.

Age presented the greatest C-statistic for the prediction of death and SARS, followed by the presence of symptoms, such as dyspnea, fever and cough, and the presence of comorbidities, like heart disease and diabetes.

### Geospatial distribution

The density of COVID-19 cases was plotted to visualize the concentration of cases, alongside the incidence, which considers the total population of each area (Fig 3). It is possible to note on Figure 3 that disease severity was not correlated with the number of cases.

**Fig 3.**
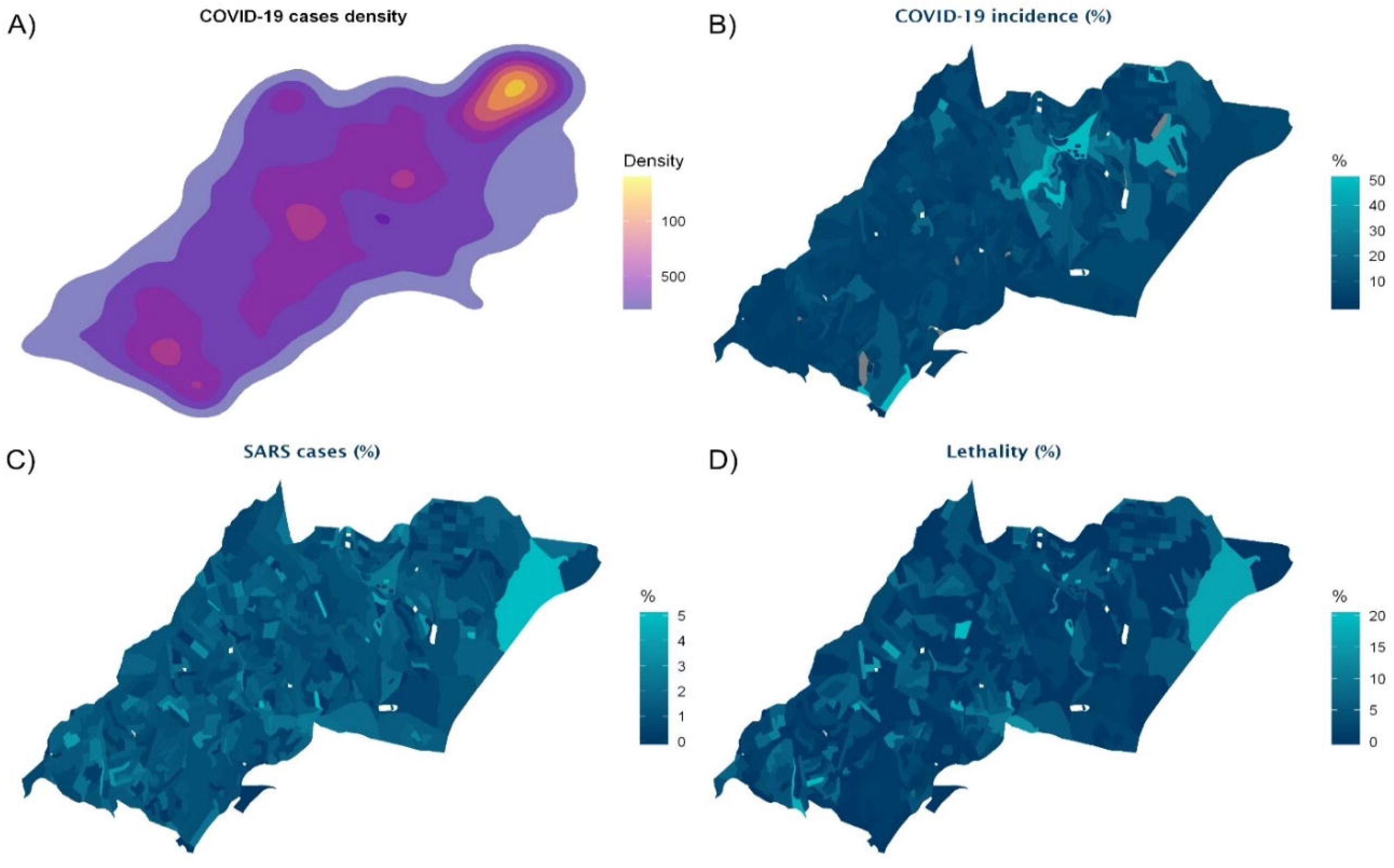
Geospatial distribution of cases and outcomes. Density of positive COVID-19 cases in the region (A), incidence of cases in the population (B), SARS incidence among positive cases (C), mortality among positive cases (D), by census sector, according to the place of residence of cases.

In map “A”, Fig 3, there is clearly an area of higher density of positive COVID-19 cases, which overlaps with the area of the largest slums in the city of São Paulo, Paraisópolis, with a predominantly young and vulnerable population living in overcrowded households. However, the incidence of positive COVID-19 cases, SARS and deaths in this area are not high, when compared to other areas in the region (Figs “3B”, “3C” and “3D”).

The data on progression to SARS was reviewed for the 16,061 individuals included in the study. After excluding individuals with missing information on comorbidities, the multiple model was adjusted based on data from 15,235 patients.

In the final multiple model (Table 2), clinical and demographic variables were more relevant than the region *per se*, which was not included in the final model, even dough place of residence had a p value <0.001, C-statistic of 0.57, and 0.74% of deviance explained in the simple analysis (Fig 4). The comorbidities found to be associated with a higher risk of SARS were obesity and immunodepression. No socioeconomic variable remained significant to be included in the multiple model. The complete model presented a C-statistic of 0.96 and deviance explained of 50%.

**Table 2.** Results of the multiple model for SARS associated with COVID-19. N=15,235.

| Factors | OR (95%CI) | p value |
| --- | --- | --- |
| Age (years) | 1.063 (1.057;1.069) | <0.001 |
| Sex (Male) | 1.61 (1.36;1.91) | <0.001 |
| Obesity (y) | 25.32 (16.31;39.31) | <0.001 |
| Immunodepression (y) | 12.15 (7.62;19.37) | <0.001 |
| Renal disease (y) | 10.12 (5.81;17.62) | <0.001 |
| Heart disease (y) | 9.79 (8.21;11.68) | <0.001 |
| Respiratory disease (y) | 6.72 (4.81;9.40) | <0.001 |
| Diabetes (y) | 6.70 (5.51;8.15) | <0.001 |
| s (Longitude, Latitude) | Fig 4 | 0.721 |

**Fig 4.**
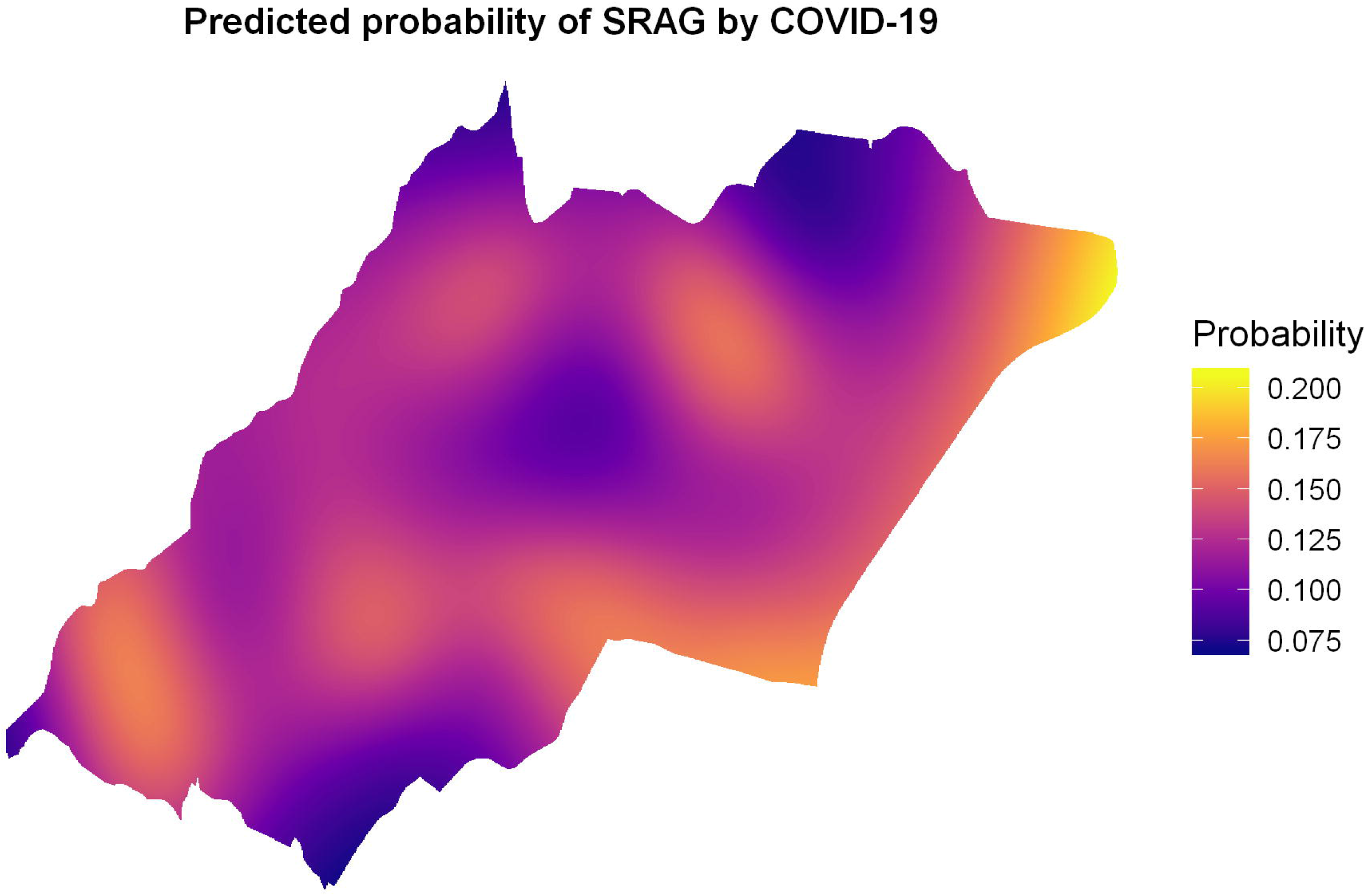
Predicted probability of progression to SARS by the space-only model. N=16,601.

The data of 15,795 individuals was reviewed for deaths. After excluding patients with no comorbidity data, the multiple model for death was adjusted based on information from 15,100 patients.

As was the case for SARS, in the final multiple model (Table 3), clinical and demographic variables were more relevant than the region *per se*, which was not included in the final model, even dough place of residence had a p value 0.001, C-statistic of 0.62, and 1.8% of deviance explained in the simple analysis (Fig 5). The comorbidities associated with a higher mortality risk were renal diseases and obesity. The complete model presented a C-statistic of 0.96 and deviance explained of 46.5%.

**Table 3.** Results of the multiple model for deaths associated with COVID-19. N=15,100.

| Factors | OR (CI 95%) | p value |
| --- | --- | --- |
| Age (years) | 1.082 (1.071; 1.092) | <0.001 |
| Sex (Male) | 1.68 (1.27; 2.21) | <0.001 |
| Renal disease (y) | 11.8 (7.1; 19.7) | <0.001 |
| Obesity (y) | 8.49 (5.21; 13.85) | <0.001 |
| Immunodepression (y) | 7.68 (4.22; 13.97) | <0.001 |
| Heart disease (y) | 4.41 (3.26; 5.97) | <0.001 |
| Respiratory disease (y) | 3.50 (2.23; 5.51) | <0.001 |
| Diabetes (y) | 3.06 (2.29; 4.09) | <0.001 |
| GeoSES (0.1 unit) | 0.896 (0.844; 0.951) | <0.001 |
| s (Longitude, Latitude) | Fig 5 | 0.657 |

**Fig 5.**
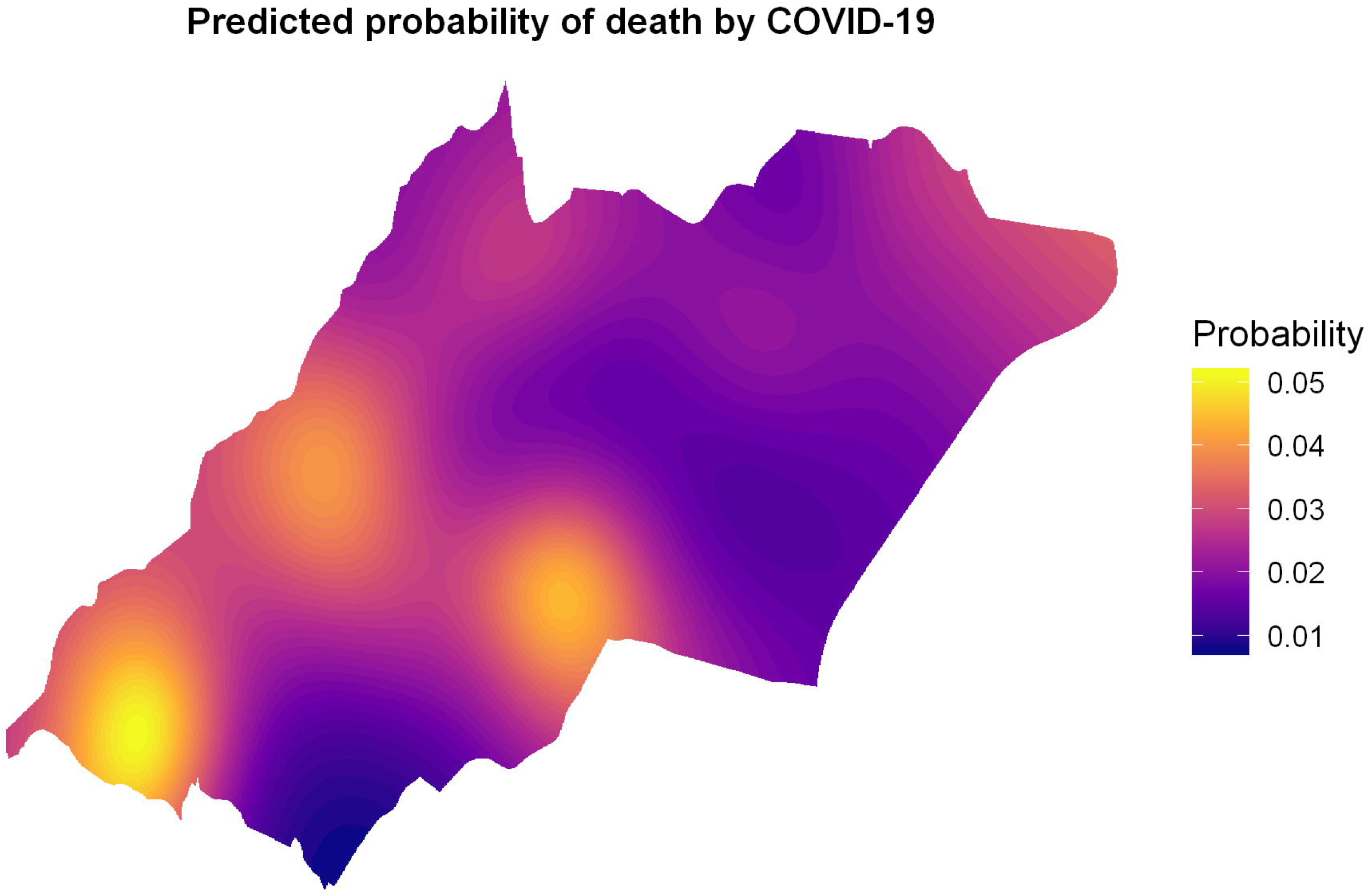
Predicted probability of death (case fatality rate/100) by the space-only model. N=15,795.

### Predictive model for death from COVID-19

The development of the predictive model included 9,931 observations. Socioeconomic and geolocation data were not included, since this information is more difficult to obtain during the patient’s clinical evaluation, but symptoms were included. The final predictive model included the following variables with their respective coefficients:

Risk score = −10.37 + (0.075 × age in years) + (0.98 × diabetes) + (1.21 × obesity) + (1.89 × immunodepression) + (0.99 × heart disease) + (2.95 × chronic renal disease) + (1.18 × respiratory disease) + (0.78 × fever) + (1.81 × dyspnea), where variables are coded as 1 when the condition is present, and 0 when absent.

Based on this formula, it is possible to calculate the predicted probability as follows:

Predicted risk (or probability) = 1 / (1+ e^− Risk score^) A risk calculator was developed, and it is accessible at: https://redcap.einstein.br/surveys/?s=PFLPC9JJ8F

To set a cutoff with a better balance between sensitivity, specificity and accuracy, the ROC curve was used to obtain a 0.0186 predicted probability with 92.2% specificity, 94.1% sensitivity and 92.2% accuracy. The C-statistic observed was 0.974 (95% CI: 0.967–0.981), demonstrating an excellent discrimination.

The training sample included 170 events, which is sufficient for the estimation of the coefficients of the 9 variables included in the final model. No collinearity between variables was evident, with the variance inflation factors ranging from 1.01 to 1.17. The maximum values for the estimated coefficient standard errors were 0.48 for the intercept, and 0.38 for immunodepression. The p value for the Hosmer-Lemeshow test was 0.485 indicating good model fit.

For internal validation, a sample of 4,966 observations was used, in which 92 events occurred (10.2 events per variable). In this sample, by applying the predicted model and maintaining the same cutoff, 92.2% sensitivity, 98.9% specificity and 92.3% accuracy was achieved. The C-statistic observed was 0.985 (95%CI: 0.978 - 0.991), with the same discriminatory quality.

For temporal validation, the predicted model for death was applied to a sample of 3,798 COVID-19 cases confirmed by PCR, notified between March and April 2021. 104 died from COVID-19 during the study period. The predictive analysis resulted in an accuracy of 80.04%, sensitivity of 98.08% and specificity of 79.53%. The C-statistic was 93.62 (95% CI: 92.24–95.00).

## Discussion

This study analyzed the space distribution and factors associated with the risk of SARS and death in positive COVID-19 cases and, derived a death risk calculator.

The risk factors associated with severe acute respiratory syndrome (SARS) were age, obesity and immunodepression, a similar finding to those of systematic reviews that evaluated risk factors associated with severe COVID-19 [21–25]. However, none of these reviews evaluated the association of environmental factors, such as living in slums, overcrowded households or in precarious sanitary conditions, with COVID-19 severity. Some observational studies have demonstrated a higher death prevalence among vulnerable and lower-income populations, but there is no solid evidence for this association [6,26].

This study found a strong association between comorbidities and age with severe COVID-19, raising the question if the poor management of chronic conditions in a vulnerable population may have contributed to the severity of COVID-19. However, no significant association was found for living conditions, while regions with a lower prevalence of chronic diseases, even the highly vulnerable ones, had a lower risk of severe respiratory syndromes.

Another key finding of this study was the increased risk of death associated with (in order of importance) age, comorbidities, and vulnerability status. The comorbidities most often related to the risk of death, in decreasing order of association, were renal disease, obesity, immunodepression, heart disease, respiratory disease, diabetes, male sex and age. Better socioeconomic conditions had a protective effect on the risk of death.

The density, incidence, severity, and case fatality rate of COVID-19 in the study region suggested the virus could spread in both high- and low-vulnerable regions. It is important to consider the vulnerability of specific groups in order to understand the impact of the pandemic on the study region, part of the largest Brazilian city with 400 thousand inhabitants, and presenting a diverse range of urban social vulnerability scores [27].

In Paraisópolis, the largest slum of the study region, there was an evident higher density of cases, in agreement with other Brazilian studies that found greater virus spread in highly vulnerable and densely populated locations [28,29]. However, a high incidence of severe cases and deaths was found in two areas of low vulnerability also presenting low density and overall incidence of cases, which is in accordance with the findings of the study by Bermudi *et al*. [29], showing high mortality in the richest areas of the city of São Paulo.

The finding of increased mortality indicate that efforts aimed at the management of chronic conditions and the care of the elderly should be the focus of public policies towards COVID-19 [30], for areas with high and low socioeconomic status.

Despite the extreme plausibility of the fact that poverty aggravates health conditions [31], this does not change the need for controlling risk factors that could be more easily modified, as well as seek strategies that would facilitate the implantation of social distancing for the most vulnerable. In Brazil, the SUS provides free and universal access to health services, with a large primary health care network present throughout the country which should be used as the transforming agents of this reality [32,33], given that countries without universal health care, such as the USA, suffered with high mortality rates from COVID-19 [34].

Predictive models for COVID-19 severe disease should include symptoms to help with medical decision making, resource planning and improvements in monitoring of COVID-19 patients [35] (i.e., the number of symptoms is predictive of disease duration [36,37]). This study identified cough, fever, sore throat, dyspnea, anosmia, loss of taste, diarrhea and fatigue as symptoms significantly associated with COVID-19 SARS and death.

The proposed risk model for death had good performance, including a C-statistic of 97.4% (95% CI: 96.7%–98.1%), which is higher than the value found by another calculator for a population of elders in the USA, with a C-statistic of 85.3%, while needing more variables [38]. Booth *et al*. [39] proposed a different strategy to predict deaths associated with COVID-19 employing molecular biomarkers measured in laboratory samples used for PCR tests; however, this approach presents limitations related to cost and availability of results in a timely manner, in addition a lower performance when compared to the model proposed by this study, with a C-statistic of 93%, sensitivity of 91% and specificity of 91%. Additional studies have focused only on the elderly or hospitalized individuals [40,41].

The model proposed by this study is a powerful response to support managers and other professionals in planning patient care, prioritizing more targeted and assertive strategies and actions, such as monitoring of positive cases with higher prediction of death, stewardship, and distribution of resources at different care levels, regions and cities, including different vulnerability contexts.

Added to the risk factors highlighted in this study, many countries in the world are going through an economic, political and ethical crisis, including Brazil, with defective response, policies that go against social distancing, lack of country-level coordination, negationism, and neoliberal policies [42], which may affect the outcomes of this pandemic. Thus, the response to the pandemic in these settings must be challenged, and larger studies including these variables in the analyses are required, such as the need for interdisciplinary analyses [43–45], considering the clinical, demographic, socioeconomic and geospatial dimensions, and incorporating all of them into one single model.

This study had the strength, of using data derived from official databases of confirmed COVID-19 cases making it possible to include a large sample size, but at the same time is subject to the limitations of observational studies and of routinely-collected health data, with the possibility of the interference by multiple errors and biases (e.g., data linkage problems, misclassification bias and underreporting) During the study period the main SARS-CoV-2 variant in circulation changed, with the introduction of the P1 variant in January of 2021, in addition to the start of the vaccination campaign at the same month. Because of these factors, the temporal validation was done finding some decrease in performance, but still finding and excellent discrimination.

In conclusion, this study contributed to the global effort to fight COVID-19, identifying risk factors associated with SARS and death among COVID-19 patients, and proposing a calculator for risk of death from COVID-19, that showed a satisfactory performance, both on a diverse setting including urban slums and extremely vulnerable populations.

## Data Availability

Individual participant data that underlie the results reported in this article, after de-identification, may be made available beginning 3 months and ending 5 years following article publication. Anyone who wishes to access the data, to achieve aims in the approved project, must direct a proposal to. To gain access, data requestors will need to sign a data access agreement.

## Acknowledgments

We are grateful to all health professionals involved in combating the pandemic of COVID-19, especially those from Campo Limpo and Vila Andrade who, in addition to patient care, contributed with the information used in this study.

## Supporting information

**S1 Fig. Spatial distribution of the health care facilities. S2 Text. STROBE Check-list**.

**S2 Text. STROBE Check-list**.

## Notes

### Competing Interest Statement

The authors have declared no competing interest.

### Funding Statement

No funding.

### Author Declarations

The study database was anonymized, with all identifiable patient information removed. This study was approved by the Institutional Review Boards of Hospital Israelita Albert Einstein and the Sao Paulo Health Department, protocol numbers 4.462.994 and 4.648.956, respectively.

## References

1. Map COVID-19. Johns Hopkins Coronavirus Resource Center. Available from <https://coronavirus.jhu.edu/map.html> Accessed on: 23 APR 2021.

2. Boletim Diário COVID-19. Available from: <https://www.prefeitura.sp.gov.br/cidade/secretarias/upload/saude/20210422_boletim_covid19_diario.pdf> Accessed on: 23 APR 2021.

3. Massuda A, Hone T, Leles FAG, de Castro MC, Atun R. The Brazilian health system at crossroads: progress, crisis and resilience. BMJ Global Health. 2018;3(4):e000829.

4. Melo EA, Mendonça M H M, Oliveira J R, Andrade G C L. Changes in the National Primary Care Policy: between setbacks and challenges. Health debate. 2018;42(spe 1):38–51.

5. Saini, K S et al. Mortality in patients with cancer and coronavirus disease 2019: A systematic review and pooled analysis of 52 studies. European Journal of Cancer. 2020;139:43–50.

6. Khan M M A, Khan M N, Mustagir M G, Rana J, Islam M S, Kabir M I. Effects of underlying morbidities on the occurrence of deaths in COVID-19 patients: A systematic review and meta-analysis. J Glob Health. 2020;10(2):020503.

7. Fahad A, et al. Early Report on Published Outcomes in Kidney Transplant Recipients Compared to Nontransplant Patients Infected with Coronavirus Disease 2019. Transplantation Proceedings. 2020. 52; 9:2659–2662.

8. Fadini, GP. Prevalência e impacto do diabetes entre pessoas infectadas com SARS-CoV-2. J Endocrinol Invest.2020; 43, 867–869.

9. Lau LL, Hung N, Wilson K. COVID-19 estratégias de resposta: considerando as desigualdades entre e dentro dos países. Int J Equity Health. 2020; 19, 137.

10. Horton R. Offline: COVID-19 is not a pandemic. Lancet. 2020; 26;396(10255):874.

11. Guimarães RB, Catão RC, Martinuci OS, Pugliesi EA, Matsumoto PSS. Geographical reasoning and the keys to reading Covid-19 in Brazilian territory. Advanced Studies, 34(99), 119–140.

12. Alcântara E. Investigating spatiotemporal patterns of the COVID-19 in São Paulo State, Brazil. Geospatial Health.2020; 15(2).

13. Fundação SEADE. São Paulo State Statistics Portal. Available from < https://www.seade.gov.br/seade-disponibiliza-projecoes-de-populacao-e-domicilios-ate-2050/> Accessed on: 23 APR 2021.

14. Pavarini SC, Barha EJ, Mendiondo MS, Filizola CL, Petrilli Filho JF, Santos AA. Family and social vulnerability: a study with octogenarians. Rev Lat Am Enfermagem. 2009;17(3):374–9.

15. Barrozo LV. GeoSES: A socioeconomic index for health and social research in Brazil. PLoS One. 2020;15(4):e0232074.

16. Royston P, Luas KGM, Altman D G, Vergouwe Y. Prognosis and prognostic research: Developing a prognostic model BMJ 2009; 338: b604

17. R Core Team (2019). R: A language and environment for statistical computing. R Foundation for Statistical Computing, Vienna, Austria. URL https://www.R-project.org/.

18. Wood SN. Fast stable restricted maximum likelihood and marginal likelihood estimation of semiparametric generalized linear models. Journal of the Royal Statistical Society. 2011;73(1):3–36.

19. H. Wickham. ggplot2: Elegant Graphics for Data Analysis. Springer-Verlag New York, 2016.

20. Simon Garnier (2018). viridis: Default Color Maps from ‘matplotlib’. R package version 0.5.1. https://CRAN.R-project.org/package=viridis.

21. Fadini GP, Morieri ML, Boscari F, Fioretto P, Maran A, Busetto L, et al. Newly-diagnosed diabetes and admission hyperglycemia predict COVID-19 severity by aggravating respiratory deterioration. Diabetes Res Clin Pract. 2020;168:108374.

22. Yang, J, Hu, J, Zhu, C. Obesity aggravates COVID-19: A systematic review and meta-analysis. J Med Virol. 2021; 93: 257– 261.

23. Yang, Jing et al. Prevalence of comorbidities and its effects in patients infected with SARS-CoV-2: a systematic review and meta-analysis. International Journal of Infectious Diseases, 2020; 94, 91 – 95.

24. Gao Ya. Impacts of immunosuppression and immunodeficiency on COVID-19: A systematic review and meta-analysis. Journal of Infection.2020;81, Issue 2, e93–e95.

25. Geng MJ, Wang LP, Ren X. Risk factors for developing severe COVID-19 in China: an analysis of disease surveillance data. Infect Dis Poverty.2021;48.

26. Baqui P. Ethnic and regional variations in hospital mortality from COVID-19 in Brazil: a cross-sectional observational study. The Lancet Global Health. 2020;8, e1018–e1026.

27. São Paulo, SEADE. Paulista Social Vulnerability Index version 2010. <http://ipvs.seade.gov.br/view/pdf/ipvs/principais_resultados.pdf> Accessed on: 23 APR 2021.

28. Castro MC, Kim S, Barberia L, Ribeiro AF, Gunzerda S, Ribeiro KB. Spatiotemporal pattern of COVID-19 spread in Brazil. Science. 2021;eabh1558.

29. Bermudi PMM, Lorenz C, Aguiar BS, Failla MA, Barrozo LV, Chiaravalloti-Neto F. Spatiotemporal ecological study of COVID-19 mortality in the city of São Paulo, Brazil: Shifting of the high mortality risk from areas with the best to those with the worst socioeconomic conditions. Travel Medicine and Infectious Disease.2021;39: 101945.

30. Fatima M, O’Keefe KJ, Wei W, Arshad S, Gruebner O. Geospatial Analysis of COVID-19: A Scoping Review. Int J Environ Res Public Health. 2021 Feb 27;18(5):2336.

31. Tracking Universal Health Coverage: 2017 Global Monitoring Report – The World Bank. Available from: <https://www.worldbank.org/en/topic/universalhealthcoverage/publication/tracking-universal-health-coverage-2017-global-monitoring-report> Accessed on: 23 APR 2021.

32. Mendonça MJ, Schilling C. Family Health Strategy, a strong model of Primary Health Care that brings results. Debate Health. 2018;42: 18–37.

33. Ventura, D F L, et al. Challenges of the COVID-19 pandemic: for a Brazilian research agenda on global health and sustainability. Cad. Public Health.2020;36,4.

34. Neil M Ferguson, Daniel Laydon, Gemma Nedjati-Gilani. Impact of non-pharmaceutical interventions (NPIs) to reduce COVID-19 mortality and healthcare demand. Imperial College London. 2020.

35. Sudre CH. Symptom clusters in Covid19: A potential clinical prediction tool from the COVID Symptom study app. MedRxiv 2020.06.12.20129056.

36. Surendra, H.Clinical characteristics and mortality associated with COVID-19 in Jakarta, Indonesia: A hospital-based retrospective cohort study Surendra, Henry et al. The Lancet Regional Health. 2020;9: 100108.

37. Garcia-Vidal, C. Trends in mortality of hospitalised COVID-19 patients: A single centre observational cohort study from Spain. The Lancet Regional Health.2020;3, 100041.

38. Ioannou GN, Green P, Fan VS. Development of COVIDVax Model to Estimate the Risk of SARS-CoV-2–Related Death Among 7.6 Million US Veterans for Use in Vaccination Prioritization. JAMA Netw Open. 2021;4(4):e214347.

39. Booth AL, Abels E, McCaffrey P. Development of a prognostic model for mortality in COVID-19 infection using machine learning. Mod Pathol. 2021 Mar;34(3):522–531.

40. Haimovich AD, Ravindra NG, Stoytchev S, Young HP, Wilson FP, van Dijk D, Schulz WL, Taylor RA. Development and Validation of the Quick COVID-19 Severity Index: A Prognostic Tool for Early Clinical Decompensation. Ann Emerg Med. 2020;76(4):442–453.

41. Sudre CH, Lee KA, Lochlain MN, Varsavsky T, Murray B, Graham MS et al. Symptom clusters in Covid-19: A potential clinical prediction tool from the COVID Symptom study app. MedRxiv 2020; 129056.

42. Hallal, PC. SOS Brazil: science under attack. The Lancet. 2020;397:373–374.

43. Franch-Pardo I, Napoletano BM, Rosete-Verges F, Billa L. Spatial analysis and GIS in the study of COVID-19. A review. Sci Total Environ. 2020. 15;739:140033.

44. Sisó-Almirall A, Kostov B, Mas-Heredia M, Vilanova-Rotllan S, Sequeira-Aymar E, Sans-Corrales M, Sant-Arderiu E, Cayuelas-Redondo L, Martínez-Pérez A, García-Plana N, Anguita-Guimet A, Benavent-Àreu J. Prognostic factors in Spanish COVID-19 patients: A case series from Barcelona. PLoS One. 2020;15(8):e0237960.

45. Covino M, De Matteis G, Santoro M, Sabia L, Simeoni B, Candelli M, Ojetti V, Franceschi F. Clinical characteristics and prognostic factors in COVID-19 patients aged ≥80 years. Geriatr Gerontol Int. 2020;20(7):704–708.

