## Supplementary material for "What most influences severity and death of COVID-19 patients in Brazil? Is it clinical, social, or demographic factors? An observational study": S1 Fig

### Supplement 1. Health care facilities locations

The distribution of the health care facilities, privet and public, accessed by the study population can be visualized in the Figure S1. Health care facilities were categorized as privet or public according to the National Registry of Health Care Facilities (CNES, in Portuguese), and was considered in the analyses as an indicator of health care access, since public health services are universally accessible, while only those with health insurance or high income have access to private services.

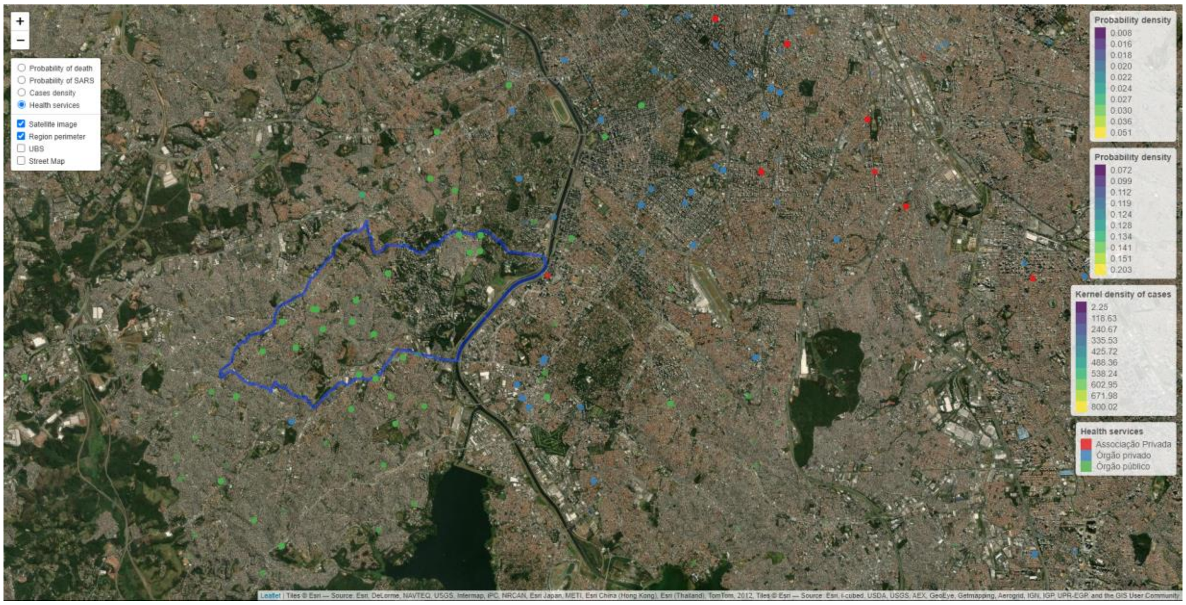


**Fig 1S. Spatial distribution of the health care facilities, privet (red and blue) and public (green), accessed by the study population.**

The map was made in the R software, version 3.6.3[1], using the leaflet[2] package.

To view the interactive version of the map, access the link:
http://apps.einstein.br/iirs/nisi/mapas/Mapas_20210413.html

It will be possible to zoom in, view the area or even select other information from the study.

### References


1. R Core Team (2019). R: A language and environment for statistical computing. R Foundation for Sta tistical Computing, Vienna, Austria. URL https://www.R-project.org/.
2. Joe Cheng, Bhaskar Karambelkar and Yihui Xie (2019). leaflet: Create Interactive Web Maps with t he JavaScript 'Leaflet' Library. R package version 2.0.3. https://CRAN.R-project.org/package=leaflet
